# COVID-19 Patients Form Memory CD8+ T Cells that Recognize a Small Set of Shared Immunodominant Epitopes in SARS-CoV-2

**DOI:** 10.1101/2020.07.24.20161653

**Authors:** Andrew P. Ferretti, Tomasz Kula, Yifan Wang, Dalena M.V. Nguyen, Adam Weinheimer, Garrett S. Dunlap, Qikai Xu, Nancy Nabilsi, Candace R. Perullo, Alexander W. Cristofaro, Holly J. Whitton, Amy Virbasius, Kenneth J. Olivier, Lyndsey B. Baiamonte, Angela T. Alistar, Eric D. Whitman, Sarah A. Bertino, Shrikanta Chattopadhyay, Gavin MacBeath

## Abstract

Development of effective strategies to detect, treat, or prevent COVID-19 requires a robust understanding of the natural immune response to SARS-CoV-2, including the cellular response mediated by T cells. We used an unbiased, genome-wide screening technology, termed T-Scan, to identify specific epitopes in SARS-CoV-2 that are recognized by the memory CD8+ T cells of 25 COVID-19 convalescent patients, focusing on epitopes presented by the six most prevalent HLA types: A*02:01, A*01:01, A*03:01, A*11:01, A*24:02, and B*07:02. For each HLA type, the patients’ T cells recognized 3–8 immunodominant epitopes that are broadly shared among patients. Remarkably, 94% of screened patients had T cells that recognized at least one of the three most dominant epitopes for a given HLA, and 53% of patients had T cells that recognized all three. Subsequent validation studies in 18 additional A*02:01 patients confirmed the presence of memory CD8+ T cells specific for the top six A*02:01 epitopes, and single-cell sequencing revealed that patients often have many different T cell clones targeting each epitope, but that the same T cell receptor Vα regions are predominantly used to recognize these epitopes, even across patients. In total, we identified 29 shared epitopes across the six HLA types studied. T cells that target most of these epitopes (27 of 29) do not cross-react with the endemic coronaviruses that cause the common cold, and the epitopes do not occur in regions with high mutational variation. Notably, only 3 of the 29 epitopes reside in the spike protein, highlighting the need to design new classes of vaccines that recapitulate natural CD8+ T cell responses to SARS-CoV-2.

## Introduction

Coronavirus Disease 2019, or COVID-19, is a global pandemic that has claimed >500,000 lives world-wide and has affected millions more. Developing effective vaccines and therapies requires understanding how the adaptive immune response recognizes and clears the virus and how the interplay between the virus and the immune system affects the pathology of the disease. To date, most efforts have focused on the B cell-mediated antibody response to the virus, but less is understood about how cytotoxic CD8+ T cells recognize and clear infected cells. Notably, the vast majority of current vaccine development efforts are focused on eliciting neutralizing antibodies to the virus, most frequently by immunizing with the spike (S) protein of SARS-CoV-2, or even with just the receptor binding domain (RBD) of the S protein *(1)*. Studies of the most closely related coronavirus, SARS-CoV, which caused the 2002/2003 outbreak of Severe Acute Respiratory Syndrome (SARS), showed that virus-specific memory CD8+ T cells persisted for six to eleven years in individuals who had recovered from SARS, whereas memory B cells and anti-viral antibodies were largely undetectable *(2, 3)*. Similarly, a recent study of COVID-19 convalescent patients showed that although antibody responses to SARS-CoV-2 could be detected in most infected individuals 10 – 15 days following symptom onset, responses declined to baseline in many patients during the study’s 3-month follow up period *(4)*. These findings suggest that vaccines focused solely on eliciting neutralizing antibodies to the S protein may be insufficient to elicit long-term immunity to coronaviruses. Notably, mouse studies of SARS-CoV showed that virus-specific CD8+ T cells are sufficient to enhance survival and diminish clinical disease *(5)* and that immunization with a single immunodominant CD8+ T cell epitope confers protection from lethal viral infection *(6)*. These studies highlight the importance of understanding the natural CD8+ T cell response to SARS-CoV-2 as a route to designing more durable vaccines.

Recently, studies using megapools of predicted T cell epitopes revealed that most COVID-19 convalescent patients, including those with severe disease, exhibit SARS-CoV-2-specific CD8+ T cells, and that at least some are directed at the S protein *(7, 8)*. To date, however, the precise targets of CD8+ T cells in convalescent patients have not been identified, and it is not known how frequently these epitopes are shared among patients, how specific they are to SARS-CoV-2, or how effectively CD8+ T cells protect against severe disease. To address these questions, we used a recently-developed high-throughput screening technology, termed T-Scan *(9)*, to simultaneously screen all the memory CD8+ T cells of 25 convalescent patients against every possible MHC Class I epitope in SARS-CoV-2, as well as SARS-CoV and the four coronaviruses that cause the common cold (HKU1, OC43, 229E, and NL63). Because T cells recognize their viral peptide targets in the context of MHC proteins, which are defined by an individual’s Human Leukocyte Antigen (HLA) type, we selected patients that are positive for each of the six most prevalent HLA types (A*02:01, A*01:01, A*03:01, A*11:01, A*24:02, and B*07:02).

Collectively, ∼90% of the U.S. population and ∼85% of the world population are positive for at least one of these six alleles *(10, 11)*. We focused our efforts on patients with relatively mild disease (primarily non-hospitalized patients) to discover the most protective epitopes, but also included patients with moderate to severe disease to determine if T cell responses correlate with disease severity. Overall, we found that the CD8+ T cell response is dominated by a few highly antigenic epitopes in SARS-CoV-2 that are shared among patients with the same HLA type. These epitopes are largely unique to SARS-CoV-2 (i.e., do not occur in “common cold” coronaviruses), are invariant among viral isolates, and are frequently targeted by multiple clonotypes within each patient. Notably, only ∼10% of the epitopes occur in the S protein. These results provide the necessary tools to better understand the CD8+ T cell response in COVID-19 and have significant implications for vaccine design and development.

## Results

We set out to determine the precise epitopes in SARS-CoV-2 that are recognized by the memory CD8+ T cells of patients that have recovered from COVID-19. To do this, we relied on a high-throughput cell-based screening technology (T-Scan) that enables simultaneous identification of the natural targets of CD8+ T cells in an unbiased, genome-wide fashion (Fig. 1A). Briefly, CD8+ T cells are co-cultured with a genome-wide library of target cells (HEK 293 cells). Each target cell in the library expresses a different 61-amino acid (61-aa) protein fragment. These fragments are processed naturally by the target cells and the appropriate peptide epitopes are displayed on class I MHC molecules on the cell surface. If a CD8+ T cell encounters its target in the co-culture, it secretes cytotoxic granules into the target cell, inducing apoptosis. Early apoptotic cells are then isolated from the co-culture and the expression cassettes are sequenced, revealing the identity of the protein fragment. Because the assay is non-competitive, hundreds to thousands of T cells can be screened against tens of thousands of targets simultaneously. To address the bottleneck of extensive sorting needed to isolate rare recognized target cells in high complexity libraries *(9)*, we engineered the target cells to express a Granzyme B (GzB)-activated version of the scramblase enzyme XKR8, which drives the rapid and efficient transfer of phosphatidylserine to the outer membrane of early apoptotic cells. Early apoptotic cells are then enriched by magnetic-activated cell sorting with Annexin V (see methods and Fig. 1A). This modification increased throughput of the T-Scan assay 20-fold, enabling the rapid processing of a large number of patient samples.

**Figure 1.**
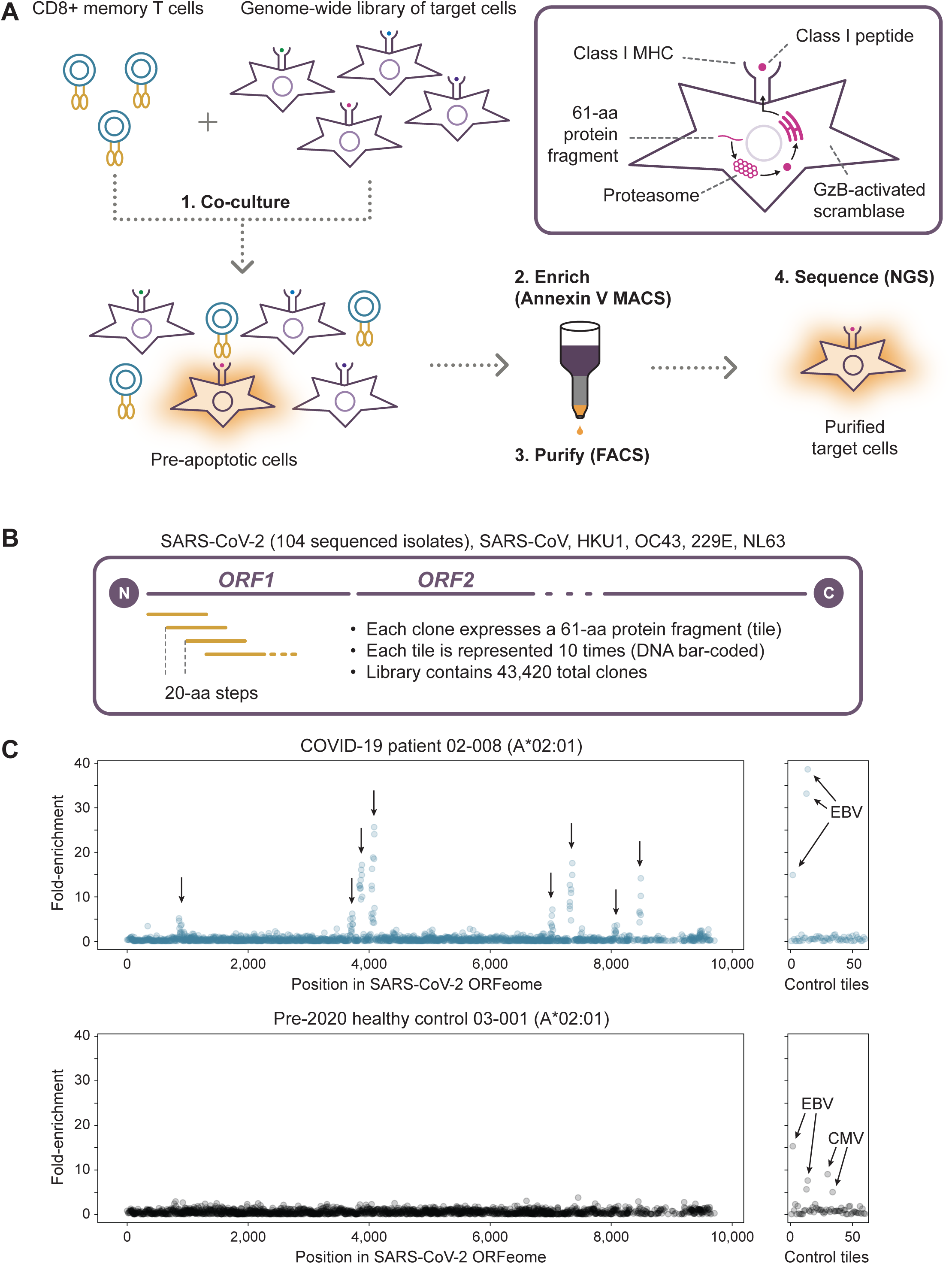
T-Scan approach for comprehensive mapping of the memory CD8+ T cell response to SARS-CoV-2. (**A**) Overview of the T-Scan antigen discovery screen. (**B**) Design of the ORFeome-wide SARS-CoV-2 antigen library. (**C**) Example SARS-CoV-2 ORFeome-wide T-Scan screen data for a convalescent COVID-19 patient (top panel) and healthy control (bottom panel). Each circle represents a single 61-aa SARS-CoV-2 protein fragment, with the X-axis showing the position of each fragment in the concatenated SARS-CoV-2 ORFeome. The Y-axis shows the performance of the fragment in the screen, calculated as the ratio of sorted target cells expressing the protein fragment relative to the unsorted target library. The right panels show the performance of the 60 positive control protein fragments derived from CMV, EBV, and Influenza.

To comprehensively map responses to SARS-CoV-2, we generated a library of 61-aa protein fragments that tiled across all 11 open reading frames (ORFs) of SARS-CoV-2 in 20-aa steps (Fig. 1B). To capture the known genetic diversity of SARS-CoV-2, we also included all protein-coding variants from the 104 isolates that had been reported as of March 15, 2020. We also included the complete set of ORFs (ORFeome) of SARS-CoV and the four endemic coronaviruses that cause the common cold (betacoronaviruses HKU1 and OC43, and alphacoronaviruses NL63 and 229E). As positive controls, we included known immunodominant antigens from Cytomegalovirus, Epstein-Barr virus, and Influenza virus *(12)*. Finally, each protein fragment was represented ten times, each encoded with a unique nucleotide barcode to provide internal replicates in our screens, for a final library size of 43,420 clones.

To understand the scope and nature of acquired immunity, we focused on the memory CD8+ T cells of convalescent COVID-19 patients. In total, we collected peripheral blood mononuclear cells (PBMCs) from 78 adult patients who had tested positive by viral PCR (swab test), had recovered from their disease, and had been out of quarantine according to Centers for Disease Control and Prevention (CDC) guidelines for at least two weeks (supplementary methods). Patients were recruited at either of two centers: Atlantic Heath System in Morristown, NJ and Ochsner Medical Center in New Orleans, LA. All patients were HLA-typed, and a summary of their characteristics are provided in table S1. As HLA A*02:01 is the most common MHC allele world-wide, we started by selecting nine HLA-A*02:01 patients with a broad range of clinical presentations: six had mild symptoms and were not hospitalized, two required supplemental oxygen, and one required invasive ventilation. In each case, we purified bulk memory CD8+ T cells (CD8+, CD45RO+, CD45RA-, CD57-) by negative selection, expanded the cells with antigen-independent stimulation (anti-CD3), and screened them against the SARS-CoV-2 library.

Target cells expressing only HLA-A*02:01 were used to provide unambiguous MHC restriction of discovered antigens. The SARS-CoV-2 screening results for one representative patient and one COVID-19-negative healthy control are shown in Figure 1C. We found reactivity to at least eight regions of SARS-CoV-2 proteins in the convalescent patient and none in the control. Importantly, we observed reproducible performance of four technical screen replicates, internal nucleic acid barcodes, and overlapping protein fragments, collectively suggesting robust screen performance. Additionally, we detected reactivity to the control CMV epitope (NLVPMVATV) in the healthy control, who was known to be CMV-positive, and reactivity to two EBV epitopes in both the COVID-19 patient and the healthy control (Fig. 1C).

Next, we examined the screen results for the full set of nine HLA-A*02:01 patients and detected reactivity to specific segments of SARS-CoV-2 ORFs in 8/9 patients (Fig. 2A). Strikingly, we found that specific fragments are recurrently recognized by the T cells of multiple patients. For example, ORF1ab aa 3881-3900 and S aa 261-280 were each recognized by 7/9 patients (Fig. 2A). Overall, we identified six regions that were targeted by CD8+ T cells from at least three different patients. In addition to being shared across patients, these regions were among the strongest responses observed in each patient. This result raises the intriguing possibility that the CD8+ T cell response to SARS-CoV-2 is largely shaped by a limited number of recurrently targeted, immunodominant epitopes.

**Figure 2.**
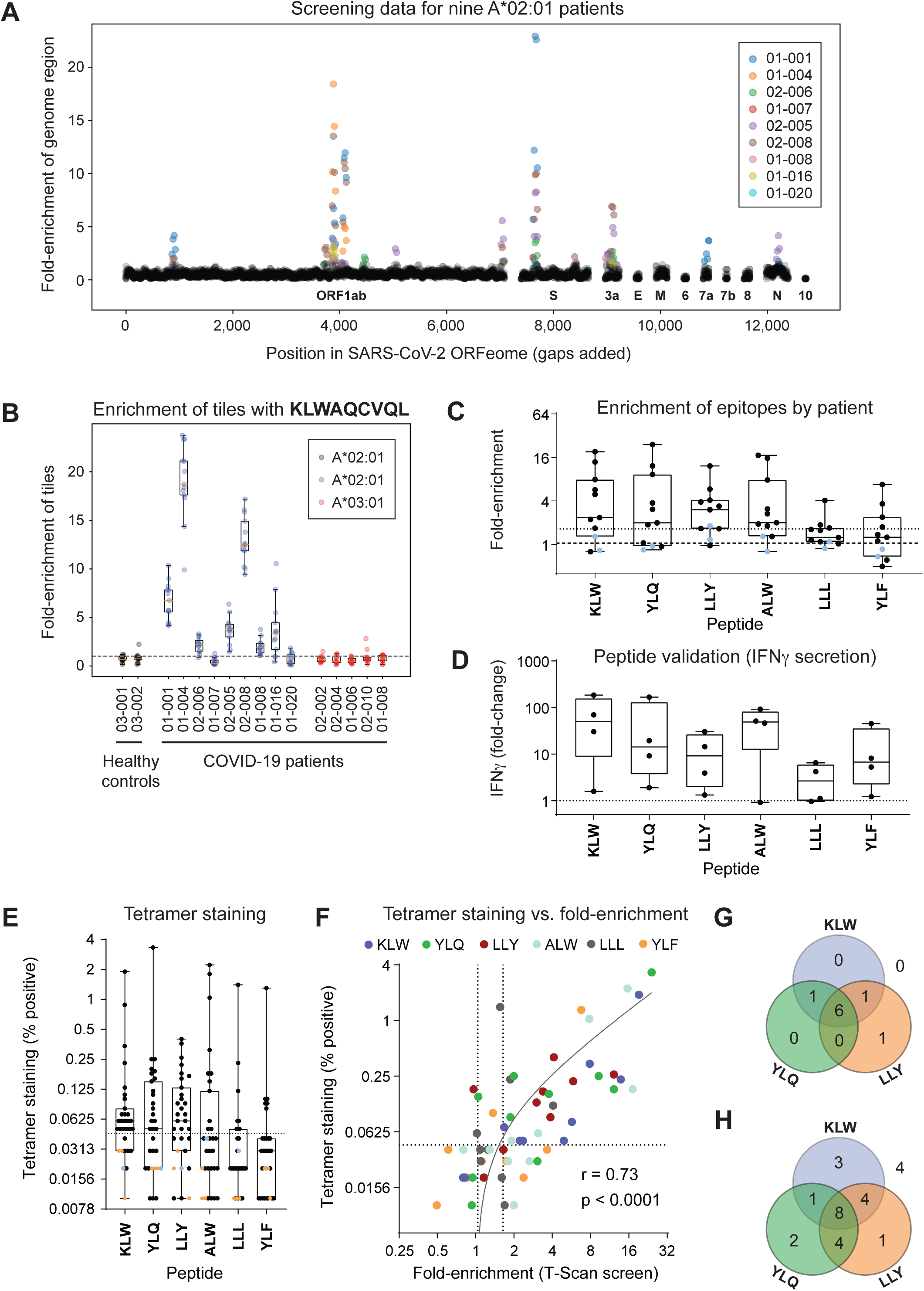
Discovery and validation of immunodominant SARS-CoV-2 epitopes presented on HLA-A*02:01. (**A**) T-Scan screen data for nine HLA-A*02:01 COVID-19 patients. Each circle corresponds to a 20aa stretch of the SARS-CoV-2 ORFeome, with the X-axis indicating the position of the stretch in the SARS-CoV-2 genome. The Y-axis shows the mean performance of all library fragments spanning the given 20aa stretch, calculated as described in Fig. 1C. Results for each patient are denoted with different colors. (**B**) Screen data for identified KLW epitope (KLWAQCVQL). The boxplots represent the screen enrichments of all fragments in the library that contain the KLW epitope. Data for the nine HLA-A*02:01 COVID-19 patient screens are shown in blue, two healthy control HLA-A*0201 screens are shown in grey, and five HLA-A*03:01 COVID-19 patient screens are shown in red. (**C**) Collapsed screen data for six identified shared epitopes. Each boxplot shows the aggregate enrichment of one epitope in each of the nine screened HLA-A*02:01 COVID-19 patients (black dots) and two healthy controls (blue dots). The Y-axis shows the mean enrichment of all fragments in the library containing the given epitope. (**D**) IFNγ ELISA validation of identified epitopes. HLA-A*02:01 target cells were pulsed with 1uM peptide and incubated with memory CD8+ T cells from four HLA-A*02:01 COVID-19 patients. The Y-axis shows the concentration of IFNγ secreted by T cells from each patient (black dot) in the presence of each peptide compared to a no-peptide control. Data are the means of two technical replicates and representative of two independent experiments. Due to sample constraints, some patient samples used in IFNγ ELISA validation had not been used in T-Scan screens. (**E**) Tetramer staining of memory CD8+ T cells reactive to six shared HLA-A*02:01 epitopes. Memory CD8+ T cells from 27 HLA-A*02:01 COVID-19 patients (black dots), one healthy HLA-matched control (blue dot), and three MHC-mismatched controls (orange dots) were stained with tetramers loaded with each of the six identified epitopes. The Y-axis indicates the percentage of tetramer-positive memory CD8+ cells. (**F**) Correlation of T-Scan screen performance and cognate T cell frequency as determined by tetramer staining. Each circle indicates the performance of one epitope in one of the nine screened HLA-A*0201 COVID19 patients. The X-axis shows the aggregate performance of the epitope in the T-Scan screen, calculated as the average enrichment of all fragments containing that epitope. The Y-axis shows the frequency of tetramer-positive memory CD8+ T cells recognizing each epitope. (**G**,**H**) Recognition of the three most common HLA-A*02:01 epitopes across COVID-19 patients based on (**G**) screening data (n=9) or (**H**) tetramer staining (n=27). For (**G**), patients were considered positive for an epitope if the aggregate performance of the epitope in the screen data exceeded a set threshold (mean + 2SD of the enrichment of all of the SARS-CoV-2 fragments in the healthy controls). For (**H**), patients were considered positive for an epitope if ≥0.05% of memory CD8+ T cells were positive by tetramer staining. Patients with no detectable reactivity to any of the three epitopes (4/27) are shown outside the Venn diagram.

We next sought to identify the precise peptide epitopes underlying the shared T cell reactivities detected in our screens. The overlapping design of our antigen library allowed us to map the T cell reactivities to specific 20-aa segments. We then used the NetMHC4.0 prediction algorithm *(13, 14)* to identify high-affinity HLA-A*02:01 peptides in each pre-identified 20-aa stretch. An example of a predicted epitope and the corresponding screen data is shown in Figure 2B (additional epitopes shown in fig. S1). Notably, the fragments scoring in our screens were enriched for high-affinity HLA-binding peptides compared to the library as a whole, further verifying their biological relevance (fig. S2). To visualize the results across all nine patients, we collapsed the screening data into a single value (mean of screen replicates and redundant tiles), revealing a set of six predicted epitopes that were recurrently recognized by three or more patients (Fig. 2C, Table 1). We then synthesized peptides corresponding to each predicted epitope to validate our findings. All six epitopes induced peptide-dependent T-cell activation as determined by interferon-gamma (IFNγ) secretion (Fig. 2D) and CD137 upregulation (fig. S3). Both IFNγ secretion and CD137 upregulation correlate with the fold enrichment in the T-Scan screen (fig. S3 and S4). As further validation, we constructed MHC tetramers with the six peptides and used them to stain the memory CD8+ T cells of all nine A*02:01 patients, as well as an additional test-set of 18 A*02:01 patients that had not been screened. Positive tetramer staining was observed in a subset of patients for all six peptides, including patients in the independent test-set (Fig. 2E). Notably, the magnitude of enrichment in the screens correlated well with the frequency of cognate T cells in the patient samples (r = 0.73, p < 0.0001; Fig. 2F), indicating that our screens detect the targets of T cells that are present at ≥0.1% frequency in the memory CD8+ T cell pool. Remarkably, the three most commonly recognized epitopes we discovered – KLW, YLQ, and LLY – are each recognized by 67% of the patients we screened, and all nine patients had a detectable response to at least one of the top three epitopes (Fig. 2G). A similar analysis of the tetramer staining data in all 27 A*02:01 patients showed recognition of at least one of these epitopes in 23/27 patients (85%; Fig. 2H). Taken together, our analysis of HLA-A*02:01 patients demonstrates the utility of the T-Scan approach in mapping SARS-CoV-2 T cell epitopes and reveals that patient T cells largely target a limited set of shared immunodominant epitopes.

**Table 1.**
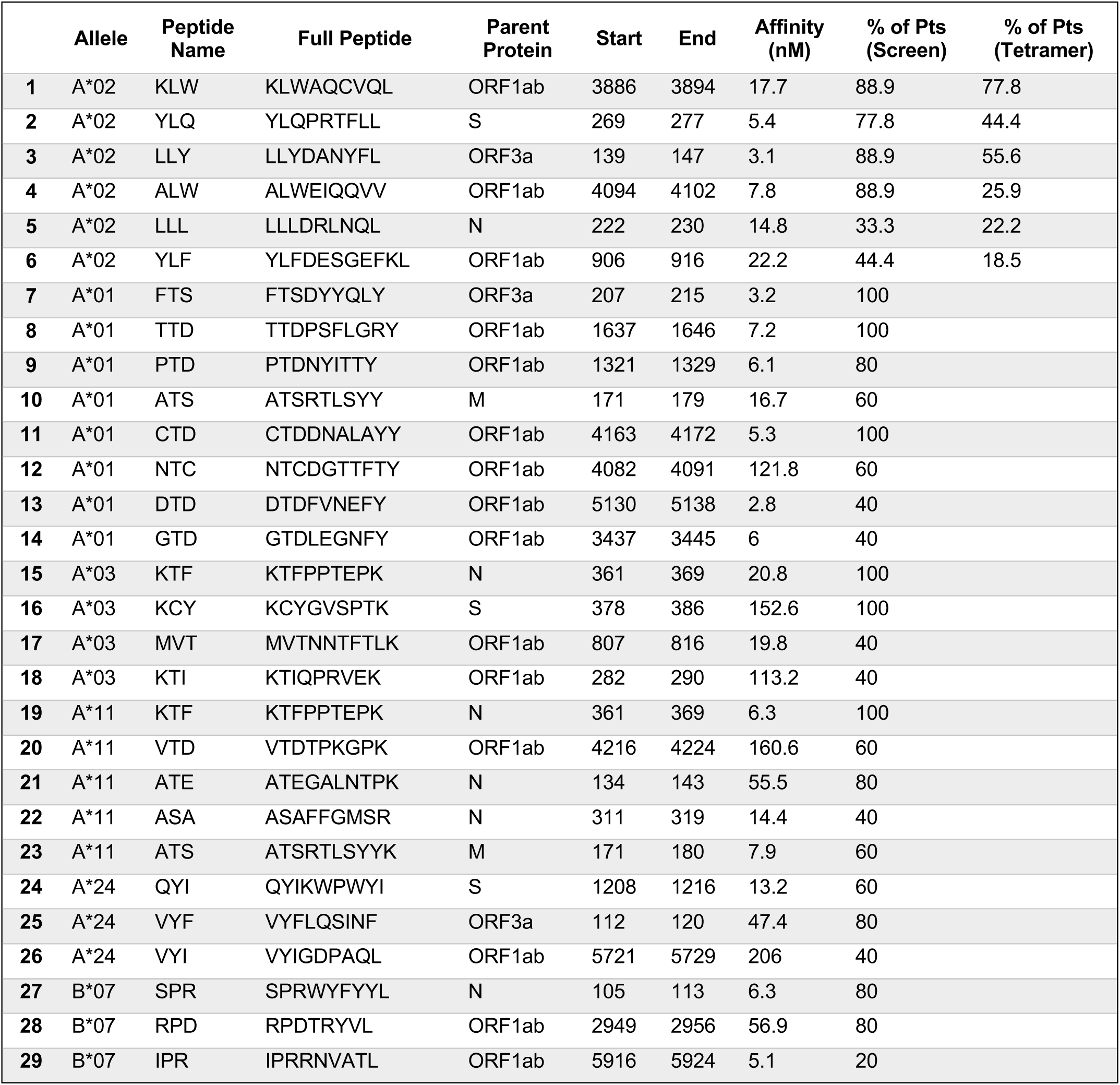
Shared immunodominant CD8+ T cell epitopes identified in convalescent COVID-19 patients.

CD8+ T cell responses are profoundly shaped by host MHC alleles, which restrict the scope of displayed peptides that serve as potential antigens. To determine whether the narrow set of immunodominant epitopes we identified for HLA-A*02:01 reflects a general feature of anti-SARS-CoV-2 CD8+ T cell responses, we mapped memory CD8+ T cell reactivities for five additional common MHC alleles: HLA-A*01:01, HLA-A*03:01, HLA-A*11:01, HLA-A*24:02, and HLA-B*07:02. Analysis of this set of HLA alleles provides a broad perspective on the nature of anti-SARS-CoV-2 CD8+ T cell immunity, as ∼90% of the U.S. population and ∼85% of the world population is positive for at least one of the six alleles we examined *(10, 11)*. For each allele, we selected five HLA+ convalescent COVID-19 patients and screened their memory CD8+ T cells against the SARS-CoV-2 library in target cells expressing only the single HLA of interest. As some patients were positive for more than one allele, their T cells were used in more than one HLA-specific screen. A total of 25 distinct patients were needed for the 34 HLA-specific screens. As with A*02:01 patients, we found robust T cell recognition of multiple regions in the SARS-CoV-2 ORFeome for patients with each HLA allele (fig. S5) and confirmed that the scoring fragments were enriched for predicted high-affinity MHC binders for each respective allele (fig. S2). Strikingly, we again observed recurrent recognition of specific protein fragments by most or all patients for each allele (Fig. 3A), indicating a narrow set of shared immunodominant responses. As before, we combined our screening data and NetMHC4.0 MHC binding predictions to map the precise epitopes underlying the top hits from our screens, and validated these peptides using IFNγ secretion (Fig. 3B) and CD137 upregulation (fig. S3) assays. We identified three or more recurrently recognized epitopes on each screened MHC allele and found that 92% of patients recognized at least one of the top three allele-specific epitopes (Fig. 3C). Collectively, we mapped and validated a set of 29 CD8+ T cell epitopes that were shared among COVID-19 patients with the same HLA type (Table 1). Most strikingly, we found that the CD8+ T cell response restricted by each of six common HLA alleles contains a limited number of recurrently targeted, immunodominant epitopes.

**Figure 3.**
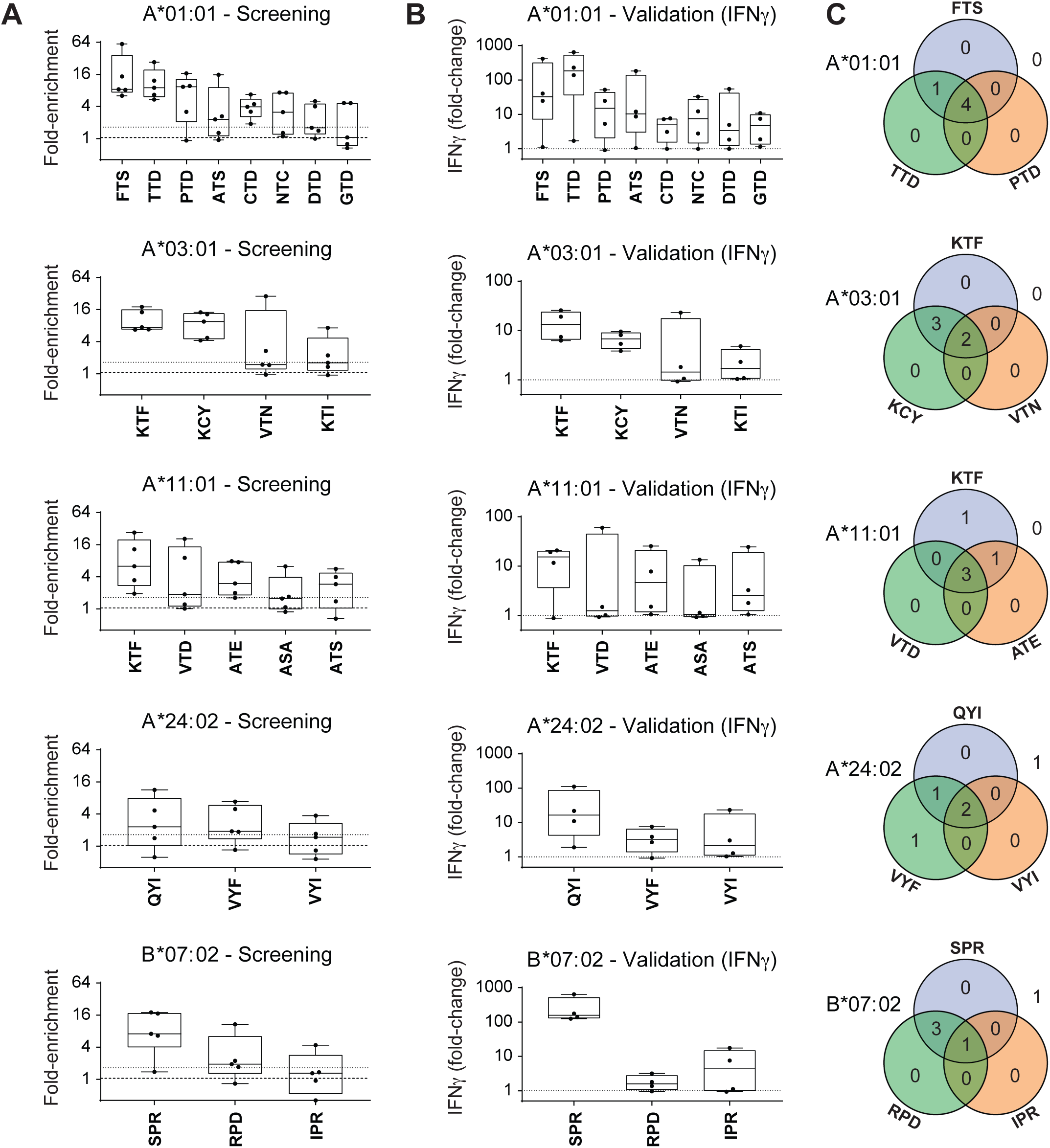
Discovery and validation of immunodominant SARS-CoV-2 epitopes presented on HLA-A*01:01, HLA-A*03:01, HLA-A*11:01, HLA-A*24:02, and HLA-B*07:02. (**A**) Collapsed T-Scan screen data for shared epitopes identified for each analyzed MHC allele. Each boxplot shows the aggregate enrichment of one epitope in each of the five COVID-19 patients (black dots) screened for the listed allele. The Y-axis shows the mean enrichment of all fragments in the library containing the given epitope. Full epitope sequences are listed in Table 1. (**B**) IFNγ ELISA validation of identified epitopes. Memory CD8+ T cells from four COVID-19 patients positive for each prioritized MHC allele were incubated with MHC-matched target cells pulsed with 1uM peptide. The Y-axis shows the concentration of IFNγ secreted by T cells from each patient (black dot) in the presence of each peptide compared to a no-peptide control. Data are the means of two technical replicates and representative of two independent experiments. (**C**) Recognition of the three most common epitopes for each prioritized MHC allele across five COVID-19 patients. Patients were considered positive for an epitope if the aggregate performance of the epitope in the screen data exceeded a set threshold (mean + 2SD of the enrichment of all of the SARS-CoV-2 fragments in the healthy controls).

The unbiased antigen mapping we performed enabled us to interrogate various features of CD8+ T cell immunity to SARS-CoV-2. First, we examined the scope of recognized viral proteins. We observed broad reactivity to many SARS-CoV-2 proteins, including ORF1ab, S, N, M, and ORF3a (Fig. 4A).

**Figure 4.**
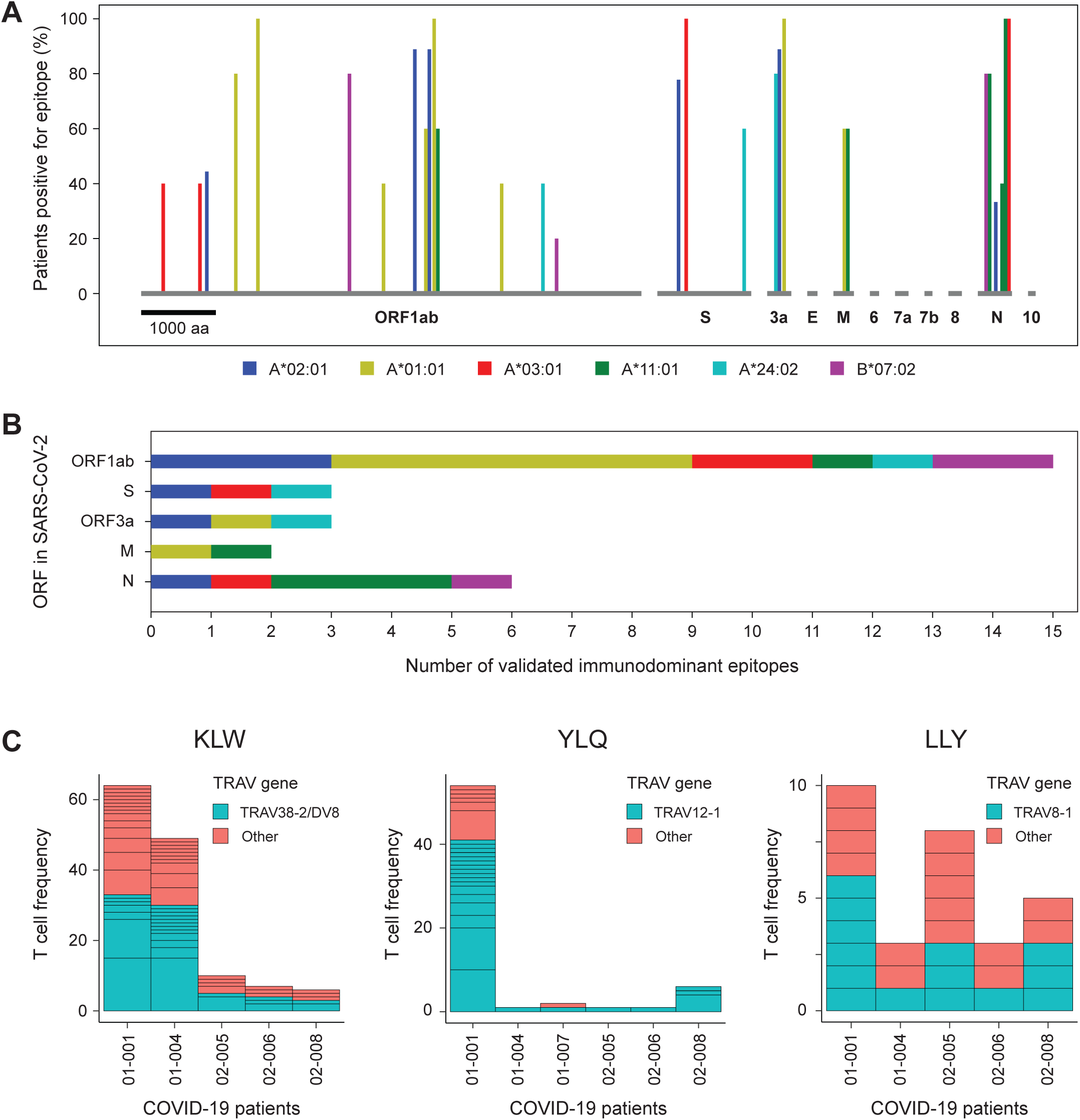
Immunodominant epitopes span the SARS-CoV-2 ORFeome and are recognized by TCRs with shared features. (**A**) Distribution of immunodominant CD8+ T cell epitopes across the SARS-CoV-2 genome. Each bar represents one validated immunodominant epitope, with the X-axis showing its position in the SARS-CoV-2 ORFeome, the color indicating its MHC restriction, and the height of the bar indicating the fraction of MHC-matched patients recognizing the epitope. Patients were considered positive for an epitope if the aggregate performance of the epitope in the screen data exceeded a threshold (mean + 2SD of the enrichment of all SARS-CoV-2 fragments in the healthy controls). For clarity, overlapping epitopes are plotted as adjacent bars. (**B**) Immunodominant CD8+ T-cell epitopes by SARS-CoV-2 ORF. The stacked bar graphs show the number of immunodominant epitopes per ORF, with the colors indicating the MHC restriction of each epitope. The MHC color-coding is the same as shown in 4a. (**C**) TCR alpha variable (TRAV) gene usage in tetramer-positive T cells across patients. Height of each box corresponds to the number of T cells within the clonotype. Blue corresponds to conserved TRAV gene for a specific epitope and red corresponds to all other TRAV genes.

Notably, only three of the 29 epitopes were located in the S protein, with most (15 of 29) located in ORF1ab and the highest density of epitopes located in the N protein (Fig. 4A,B). When taken in aggregate, our results are largely consistent with previous ORF-level analyses using peptide pools *(7, 8, 15-17)*. However, our approach provided an increased level of granularity that enabled us to identify specific epitope sequences and highlighted allele-specific differences. For example, we observed immunodominant epitopes in the S protein for HLA-A*02:01, HLA-A*03:01, and HLA-A*24:02, but not for HLA-A*01:01, HLA-A*11:01, or HLA-B*07:02. Notably, we detected only one recurrent response in the receptor-binding domain (RBD) of the S protein (KCY on HLA-A*03:01).

Next, we asked how the CD8+ T cell response to SARS-CoV-2 intersects with the emerging genetic diversity of the virus. Recent analyses, which examined the genome sequences of over 10,000 isolates of SARS-CoV-2 sampled from 68 different countries, identified a set of 28 non-synonymous coding mutations detected in at least 1% of strains *(18)*. Only one of these mutations (M protein T175M, detected in 2% of strains) was found in the immunodominant epitopes we identified (HLA-A*01:01 ATS and HLA-A*11:01 ATS). This suggests that the recognition of the epitopes we identified is unlikely to be significantly influenced by the SARS-CoV-2 genetic diversity observed thus far.

Identifying specific SARS-CoV-2 epitopes allowed us to examine the features of the T cell receptors (TCRs) recognizing these immunodominant epitopes. We used tetramers loaded with three HLA-A*02:01 epitopes (KLW, YLQ, and LLY) to stain and sort antigen-specific memory CD8+ T cells from the initial nine HLA-A*02:01-positive convalescent COVID-19 patients. We then used 10x Genomics single-cell sequencing to identify the paired TCR alpha and TCR beta chains expressed by these T cells. We identified paired clonotypes reactive to each antigen in 5/9 (KLW, ALW) or 6/9 (YLQ) patients. For a majority of responses (9/16), we detected oligoclonal recognition by five or more distinct clonotypes. Next, we examined the TCR sequences themselves. We observed striking similarity among the TCRs recognizing each antigen in terms of Vα gene segment usage and, to a lesser extent, Vβ usage (Fig. 4C). Specifically, 26/61 KLW-reactive clonotypes used TRAV38-2/DV8, 24/31 YLQ-reactive clonotypes used TRAV12-1, and 14/29 LLY-reactive clonotypes used TRAV8-1. Notably, these dominant Vα genes were used across all of the patients for whom we identified reactive clonotypes. Taken together, these data suggest that the epitopes we identified are recognized by TCRs with shared sequence features and raise the possibility that their immunodominance is shaped by the structural requirements for high-affinity TCR binding to these peptide-MHC complexes.

Another key question is how pre-existing immunity to other coronaviruses shapes the CD8+ T cell response to SARS-CoV-2. There are four commonly circulating coronaviruses, OC43, HKU1, NL63, and 229E, and cross-reactive responses to these viruses have been theorized as a potential protective factor during SARS-CoV-2 infection *(19)*. Moreover, understanding the extent of cross-reactivity has implications for accurately monitoring T cell responses to SARS-CoV-2 and for optimizing vaccine design. If the immune response to SARS-CoV-2 is shaped by pre-existing CD8 T cells that recognize other coronaviruses, we reasoned that COVID-19 patients should have reactivity to the regions of the other coronaviruses that correspond to the SARS-CoV-2 immunodominant epitopes we identified. We therefore examined T-cell reactivity to SARS-CoV-2, SARS-CoV, and all four endemic coronaviruses in the 34 genome-wide screens that we conducted across all patients and MHC alleles (Fig. 5A). We observed broad reactivity to the corresponding epitopes in SARS-CoV in over half of cases, consistent with a recent study reporting the existence of long-lasting memory T cells cross-reactive to SARS-CoV-2 in patients that had been infected in SARS-CoV during the 2002/2003 SARS outbreak *(8)*. In contrast, however, we detected almost no reactivity to OC43 and HKU1 (2/29 dominant epitopes) and none to NL63 and 229E. Beyond the 29 epitopes, we observed no reproducible cross-reactivity to any other regions of the four endemic coronaviruses, again suggesting that prior exposure to these viruses is unlikely to provide T cell-based protection from SARS-CoV-2.

**Figure 5.**
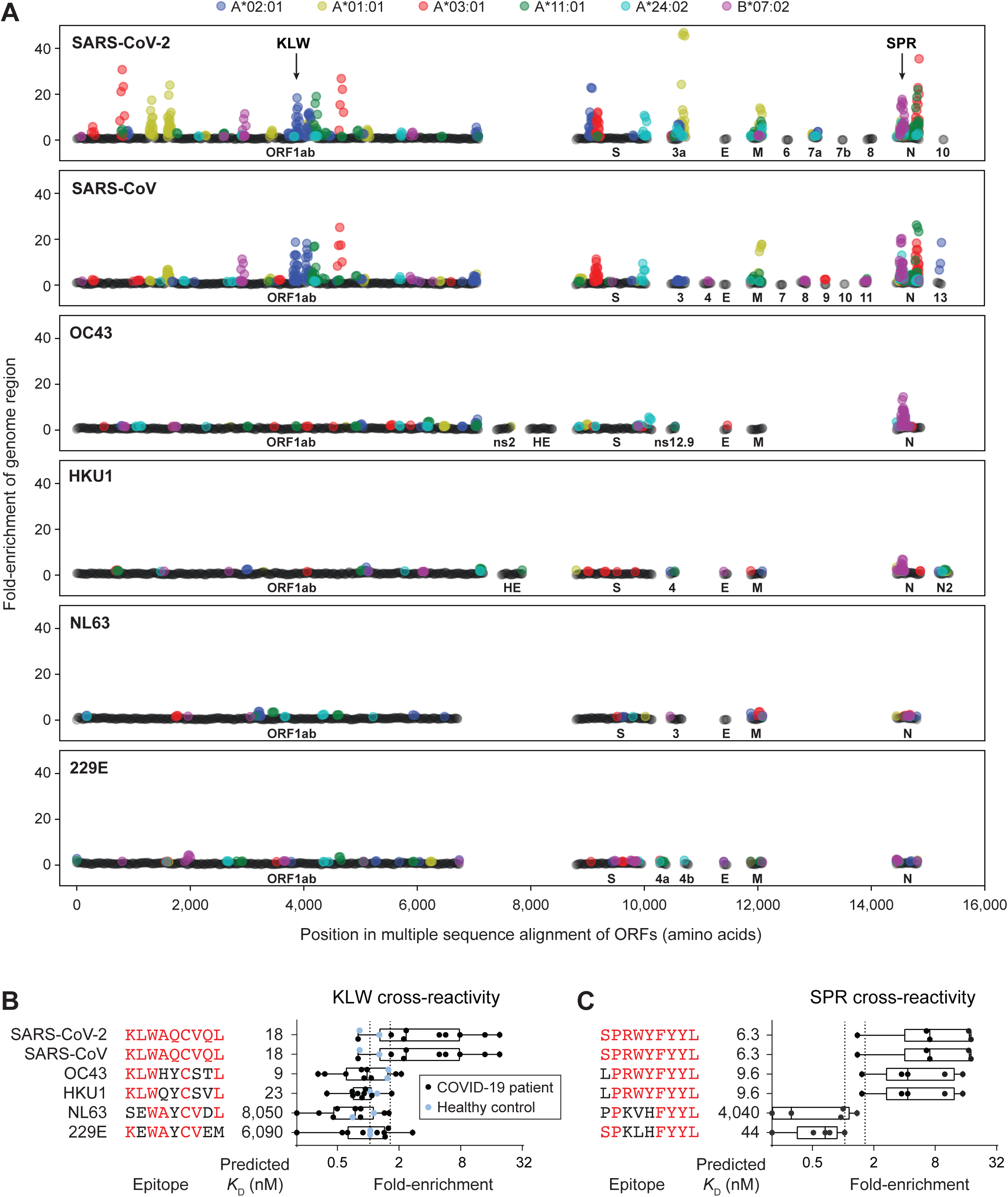
Minimal cross-reactivity of SARS-CoV-2-reactive memory CD8+ T cells with other coronaviruses. (**A**) Screen data compared across coronavirus ORFeomes. Each panel shows the collective reactivity to one coronavirus genome (SARS-CoV-2, SARS-CoV-1, OC43, HKU1, NL63, or 229E) detected in the 34 T-Scan screens performed. Each circle corresponds to a 20-aa stretch of the coronavirus ORFeome, with the X-axis indicating the position of the stretch in the ORFeome. The Y-axis shows the mean performance of all of the library fragments spanning the given 20-aa stretch, calculated as described in Fig. 1C. Results include nine HLA-A*02:01 patients, five HLA-A*03:01 patients, five HLA-A*01:01 patients, five HLA-A*11:01 patients, five HLA-A*24:02 patients, and five HLA-B*07:02 patients. For visualization, the positions of the conserved ORF1ab, S, M, E, and N proteins were aligned across all ORFeomes. (**B**) Alignment of the KLW epitope across coronavirus genomes. The alignment shows the region of each coronavirus genome corresponding to the SARS-CoV-2 HLA-A*02:01 KLW epitope. The boxplots show the aggregate screen performance of all fragments containing each epitope variant for nine HLA-A*02:01-positive COVID-19 patients (black dots) and two HLA-A*02:01-positive healthy controls (blue dots). (**C**) Alignment of the SPR epitope across coronavirus genomes. The alignment shows the region of each coronavirus genome corresponding to the SARS-CoV-2 HLA-B*07:02 epitope. The boxplots show the aggregate screen performance of all fragments containing each epitope variant for five HLA-B*07:02-positive COVID-19 patients (black dots).

Mapping the specific immunodominant epitopes in SARS-CoV-2 enabled us to determine the molecular basis for this lack of cross-reactivity. In some cases, the corresponding region is poorly conserved in the other coronaviruses and high-affinity binding to MHC is lost (see, for example, the corresponding regions of the KLW epitope in NL63 and 229E; Fig. 5B). In other cases, the corresponding epitopes are still predicted to bind with high affinity to MHC, but SARS-CoV-2-reactive T cells do not recognize them (see, for example, the corresponding regions of the KLW epitope in OC43 and HKU1; Fig. 5B).

In one notable case, we did identify a strong cross-reactive response. The HLA B*07:02 epitope SPR, which lies in the N protein, is highly conserved across betacoronaviruses and all four of the patients that demonstrated reactivity to SPR also exhibited reactivity to the corresponding epitopes in OC43 and HKU1 (Fig. 5C). Overall, however, we conclude that the CD8+ T cell response to SARS-CoV-2 is not significantly shaped by pre-existing immunity to endemic coronaviruses.

Although our study was not designed to test specific clinical hypotheses, we looked for associations between SARS-CoV-2-reactive T cells (frequency based on tetramer staining, total reactivity based on screen data) and clinical characteristics (fig. S6). No obvious associations were observed with age, sex, or disease severity, but significant negative correlations were observed with time from diagnosis to blood draw (p=0.015 for screening data and p=0.0012 for tetramer staining data; fig S6A). This is expected, as anti-viral T cells, including effector memory cells, naturally contract following an acute infection *(20, 21)*. We were particularly interested in determining if T cells specific for immunodominant epitopes confer protection from severe disease, but the number of hospitalized patients and, in particular, the number requiring invasive ventilation, was insufficient to answer this question. Appropriately powered studies to address this question are warranted but should be careful to control or adjust for time from diagnosis to blood collection.

## Discussion

To understand the natural CD8+ T cell response to SARS-CoV-2, we used an unbiased, genome-wide method that enables identification of the precise epitopes presented on MHC and functionally recognized by memory CD8+ T cells in convalescent patient blood. All 29 epitopes identified in this study were validated using independent functional assays, and the A*02:01-restricted epitopes were further validated in an independent test set of 18 patients. Overall, we found a core set of 3-8 immunodominant epitopes for each MHC allele we studied. These epitopes were recurrently targeted across patients, but also represented the strongest hits in our screens within each patient, indicating that they are both shared and dominant. Moreover, these epitopes are almost entirely specific to SARS-CoV-2/SARS-CoV, suggesting that the T cell response to SARS-CoV-2 is not significantly shaped by pre-existing immunity to the four endemic coronaviruses that cause the common cold.

Our results contrast with *in silico* studies predicting epitopes presented by HLA alleles. For example, hundreds of SARS-CoV-2-derived peptides are predicted to bind with high affinity to HLA-A*02:01 *(22)*, yet our analysis of actual T-cell responses reveals eight or fewer dominant A*02:01-restricted targets per patient. Based on the strong correlation we observed between our screening data and tetramer staining, we estimate that our screens detect T cell specificity that is present at a frequency of ≥0.1% in the pool of memory cells. Although there are likely other virus-specific T cells below this frequency, the ones we detect represent the most expanded clones and so are likely to be most important in providing protection from future infection. Generating a T cell response depends not only on high-affinity binding of the peptide to the MHC, but also on efficient processing and loading of the peptide, as well as efficient recognition of the peptide by TCRs in the naïve repertoire of the patient. Indeed, our clonotype analysis of the three most dominant A*02:01 epitopes (KLW, YLQ, and LLY) revealed that the T cell response is oligoclonal but dominated by specific T cell receptor Vα chains that are similarly shared across patients. This highlights the importance of experimentally identifying immunodominant epitopes in an unbiased fashion.

Our study also highlighted differences across MHC alleles in the total number of recognized epitopes and the proteins in which they reside. This emphasizes the importance of searching for MHC associations with disease outcome and of detailed tracking of MHC alleles in immune monitoring of vaccine trials. Previous studies using megapools of peptides spanning each of the ORFs in SARS-CoV-2 showed CD4+ and CD8+ T cell responses in all COVID-19 convalescent patients *(7, 8, 15-17)*. Although most of the reactivity to the S protein came from CD4+ T cells, some reactivity to the S protein was also observed in CD8+ T cells. Consistent with these findings, we identified 3 immunodominant epitopes in the S protein. Overall, however, we found that 90% of the CD8+ T cell reactivity was directed at epitopes outside the S protein. Grifoni et al. also showed reactivity to the M protein *(7)*, while Le Bert, et al. found reactivity to nsp7 and nsp13, which derive from ORF1ab *(8)*. We now provide specific epitopes within these proteins, as well as their MHC restriction. In contrast to peptide pool studies that found T cells in unexposed individuals that were cross reactive to SARS-CoV-2, however, our data show that the immunodominant epitopes are largely specific for SARS-CoV-2 and are not shared with other coronaviruses. If pre-existing memory responses to other coronaviruses were able to efficiently recognize SARS-CoV-2, the reacting T cells would be expected to expand, and their targets would be detected in our screens. As a result, the paucity of cross-reactive responses we found argues against substantial protection against SARS-CoV-2 stemming from CD8+ T cell immunity to the four coronaviruses that cause the common cold.

The additional level of granularity provided by identifying the specific epitopes also provides the necessary tools for tracking SARS-CoV-2-specific CD8+ T cell responses in exposed individuals or in subjects participating in vaccine trials. Diagnosis of previous exposure to SARS-CoV-2 currently relies on serological testing for antibodies that wane with time. A recent study found that IgG responses to SARS-CoV-2 decline rapidly in >90% of infected individuals in the 2-3-month period post infection, with 40% of asymptomatic individuals turning seronegative *(23)*. Additionally, follow-up studies following the 2002/2003 SARS outbreak showed that >90% of patients had no detectable memory B cells six years later *(2, 3)*. In contrast, memory T cells to SARS-CoV-2 may persist longer, as T cells specific for SARS-CoV were detected 11 or even 17 years after the 2003 SARS outbreak *(8, 24)*. Detecting SARS-CoV-2-specific CD8+ T cells can potentially be performed at large scale using an IFNγ release assay similar to commercial assays used for tuberculosis testing *(25)*. Although the frequency of SAR-CoV-2-specific memory T cells decreases in the weeks following recovery from an acute infection, the remaining pool of memory T cells can be expanded *in vitro* by stimulation with peptide epitopes, as previously demonstrated for the detection of T cells to SARS-CoV *(8, 24)*. In contrast to serological testing for antibodies, this potentially enables a diagnostic test that can detect prior exposure to COVID-19 for a prolonged period following viral infection. These tests could also be used in future studies to determine if T cell reactivity to any or all of the immunodominant epitopes correlates with disease severity or are protective against future infection.

Our findings also have significant implications for vaccine development. A majority of the T cell responses we mapped fall outside of the S protein. Only one is in the receptor binding domain of S. This argues that more robust CD8+ T cell responses across diverse patients could be generated by incorporating additional antigens into vaccine designs. We provide specific regions of the ORF1ab protein that could be used. The smaller proteins N, M, and ORF3a also appear strongly and broadly immunogenic. The epitopes that we identified in our study carry the additional benefit that they occur in regions that have thus far been subject to minimal genetic variation. While there does not appear to be significant cross-reactivity with other coronaviruses, the few regions we identified that are highly conserved and immunogenic may be of specific interest because they may confer protection across different coronaviruses and result in more robust vaccine responses by boosting pre-existing memory CD8+ T cells. Studying these epitopes in prospective tracking studies can determine whether previous exposure to other coronaviruses elicits protective or pathological immune responses.

The finding that the immunodominant epitopes for CD8+ T cells reside largely outside the spike protein raises the possibility that many of the S protein-directed vaccines currently under development may elicit an insufficient CD8+ T cell response. It should be noted that a recent vaccine candidate, BNT162b1, an RNA vaccine encoding the receptor binding domain of the S protein, did elicit CD8+ T cell responses in 80% of participants *(26)*. Given that we observed only a single A*03:01-restricted immunodominant epitope in the RBD, it is unlikely that the responses observed in this study are all directed at this epitope. Additional immunodominant epitopes may be presented by MHC alleles we did not examine, although it is unlikely that a large number of rare alleles display RBD-derived immunodominant epitopes while the six most prevalent alleles collectively feature only one. A more likely explanation is that vaccinating with a high dose of an RNA-based vaccine encoding a single protein domain could potentially elicit CD8+ T cells that recognize subdominant epitopes. It remains to be determined if this is suitably protective or if vaccines like this would benefit from additional peptides/proteins that elicit the naturally occurring shared epitopes.

In summary, our study found that memory CD8+ T cell responses in convalescent COVID-19 patients are directed against a small set of immunodominant epitopes that are shared across the majority of patients with the same HLA types. These epitopes are largely outside the spike protein, the current target of the most advanced vaccines against SARS-CoV-2. These findings enable the development of diagnostic tests for previous exposure to SARS-CoV-2 and support the inclusion of other antigens in vaccines against this virus that are more likely to mimic the natural CD8+ T cell response to SARS-CoV-2.

## Data Availability

Analyses of all data are provided in the manuscript and supplement. Patient data and identified T cell epitopes are provided in tables.

## Ethics statements

All donors provided written consent. The study was conducted in accordance with the Declaration of Helsinki (1996), approved by the Atlantic Health System Institutional Review Board and the Ochsner Clinic Foundation Institutional Review Board and registered at clinicaltrials.gov (# NCT04397900). Details of the sample collection design and all other methods are provided in supplementary materials.

## Acknowledgements

We thank the patients and their families who participated in these studies. This work was supported by TScan Therapeutics, a privately-owned biotechnology company.

## Author contributions

A.P.F., T.K., and G.M. conceived the project. A.P.F., T.K., Y.W., D.M.V.N., A.W., G.S.D., Q.X., N.N., C.R.P., H.J.W., A.V., and G.M. designed, performed, and analyzed experiments. T.K., A.F., A.W.C., Q.X, and G.S.D. designed and performed bioinformatics analyses. L.B.B., A.T.A., and E.D.W. directed sample collection and recruited patients. S.C. designed the sample collection study. K.J.O. and S.A.B. oversaw sample collection and tracking for TScan Therapeutics. A.P.F., T.K., S.A.B., S.C., and G.M. wrote the manuscript.

## References

1. N. Vabret et al., Immunology of COVID-19: Current State of the Science. Immunity 52, 910–941 (2020).

2. F. Tang et al., Lack of peripheral memory B cell responses in recovered patients with severe acute respiratory syndrome: a six-year follow-up study. J Immunol 186, 7264–7268 (2011).

3. H. Peng et al., Long-lived memory T lymphocyte responses against SARS coronavirus nucleocapsid protein in SARS-recovered patients. Virology 351, 466–475 (2006).

4. J. Seow et al., Longitudinal evaluation and decline of antibody responses in SARS-CoV-2 infection. (2020).

5. J. Zhao, J. Zhao, S. Perlman, T cell responses are required for protection from clinical disease and for virus clearance in severe acute respiratory syndrome coronavirus-infected mice. J Virol 84, 9318–9325 (2010).

6. R. Channappanavar, C. Fett, J. Zhao, D. K. Meyerholz, S. Perlman, Virus-specific memory CD8 T cells provide substantial protection from lethal severe acute respiratory syndrome coronavirus infection. J Virol 88, 11034–11044 (2014).

7. A. Grifoni et al., Targets of T Cell Responses to SARS-CoV-2 Coronavirus in Humans with COVID-19 Disease and Unexposed Individuals. Cell, (2020).

8. N. Le Bert et al., SARS-CoV-2-specific T cell immunity in cases of COVID-19 and SARS, and uninfected controls. Nature, (2020).

9. T. Kula et al., T-Scan: A Genome-wide Method for the Systematic Discovery of T Cell Epitopes. Cell 178, 1016–1028 e1013 (2019).

10. M. Maiers, L. Gragert, W. Klitz, High-resolution HLA alleles and haplotypes in the United States population. Hum Immunol 68, 779–788 (2007).

11. F. F. Gonzalez-Galarza et al., Allele frequency net database (AFND) 2020 update: gold-standard data classification, open access genotype data and new query tools. Nucleic Acids Res 48, D783–D788 (2020).

12. J. R. Currier et al., A panel of MHC class I restricted viral peptides for use as a quality control for vaccine trial ELISPOT assays. Journal of immunological methods 260, 157–172 (2002).

13. M. Andreatta, M. Nielsen, Gapped sequence alignment using artificial neural networks: application to the MHC class I system. Bioinformatics 32, 511–517 (2016).

14. M. Nielsen et al., Reliable prediction of T-cell epitopes using neural networks with novel sequence representations. Protein Sci 12, 1007–1017 (2003).

15. J. Braun et al., Presence of SARS-CoV-2 reactive T cells in COVID-19 patients and healthy donors. medRxiv, (2020).

16. C. Thieme et al., The SARS-CoV-2 T-cell immunity is directed against the spike, membrane, and nucleocapsid protein and associated with COVID 19 severity. (2020).

17. D. M. Altmann, R. J. Boyton, SARS-CoV-2 T cell immunity: Specificity, function, durability, and role in protection. Science Immunology 5, (2020).

18. T. Koyama, D. Platt, L. Parida, Variant analysis of SARS-CoV-2 genomes. Bulletin of the World Health Organization 98, (2020).

19. J. Cui, F. Li, Z. L. Shi, Origin and evolution of pathogenic coronaviruses. Nat Rev Microbiol 17, 181–192 (2019).

20. V. P. Badovinac, B. B. Porter, J. T. Harty, Programmed contraction of CD8+ T cells after infection. Nature immunology 3, 619–626 (2002).

21. E. J. Wherry, R. Ahmed, Memory CD8 T-cell differentiation during viral infection. Journal of virology 78, 5535–5545 (2004).

22. A. Nguyen et al., Human leukocyte antigen susceptibility map for SARS-CoV-2. Journal of virology, (2020).

23. Q.-X. Long et al., Clinical and immunological assessment of asymptomatic SARS-CoV-2 infections. Nature medicine, 1–5 (2020).

24. O. W. Ng et al., Memory T cell responses targeting the SARS coronavirus persist up to 11 years post-infection. Vaccine 34, 2008–2014 (2016).

25. C. Albert-Vega et al., Immune Functional Assays, From Custom to Standardized Tests for Precision Medicine. Front Immunol 9, 2367 (2018).

26. M. J. Mulligan et al., Phase 1/2 study to describe the safety and immunogenicity of a COVID-19 RNA vaccine candidate (BNT162b1) in adults 18 to 55 years of age: interim report. medRxiv, (2020).

